# Self-perceived communication competence of adults who stutter following Communication-Centered Treatment (CCT)

**DOI:** 10.1101/2023.06.26.23291589

**Authors:** Geoffrey A. Coalson, Danielle Werle, Robyn Croft, Courtney T. Byrd

## Abstract

**Purpose:** The purpose of this study was to assess self-perceived communication competence of adults who stutter following a unique treatment program – Communication-Centered Treatment (CCT) – that focuses on communication competence as one of four clinical goals of the Blank Center CARE Model^™^ (Communication, Advocacy, Resiliency, Education).

**Method:** Thirty-three adults who stutter completed the Self-Perceived Communication Competence scale (McCroskey & McCroskey, 1988) before and after their CCT program.

**Results:** Findings indicate significant gains in self-perceived communication competence post-treatment across four speaking contexts (public presentation, large meeting, small group interaction, dyadic interaction) and three audience types (strangers, acquaintances, friends). Pre-treatment stuttering frequency did not predict post-treatment gains in communication competence.

**Conclusions:** Adults who stutter consider themselves stronger communicators following a treatment designed to increase communication competence. [*ClinicalTrials.gov* NCT05908123; https://clinicaltrials.gov/show/NCT05908123]

## Introduction

Although theoretical descriptions of communication competence are not defined by fluency (e.g., Morreale et al., 2016; Spitzberg, 2013; Spitzberg & Cupach, 2011), the majority of treatment approaches for adult stuttering (e.g., O’Brian et al., 2018; Van Riper, 1973) define communication competence by increased speech fluency, with one exception. The Blank Center CARE Model^™^ (Byrd et al., 2016, 2018, 2021, 2022, 2023) trains participants in critical elements of *communication competence* in the absence of any goals designed conceal or eliminate stuttered speech. These communication competencies are then applied across a series of functional and challenging speaking scenarios with no attempts to hide or alter stuttered speech. Preliminary data found that clinicians (Byrd et al., 2021) and naïve observers (Byrd et al., 2023) rated adults who stutter as significantly stronger and more effective communicators at post-treatment compared to pre-treatment.

To date, published outcomes of communication competence following Communication-Centered Treatment (CCT) have been largely restricted to listener perspectives (Byrd et al., 2021; 2023). Although listener perception is one critical factor of effective communication, the primary clinical outcome of interest in CCT is the client’s perception of their own communication competence after treatment. To date, no study of CCT has examined self-assessed post-treatment communication competence. To investigate self-rated judgment of communication competence in the present study, adults who stutter completed the Self-Perceived Communication Competence (SPCC; McCroskey & McCroskey, 1988; Richmond & McCroskey, 1998) scale before and after CCT.

### Communication competence of people who stutter

The perception that adults who stutter are poor communicators is pervasive and has been reported across a variety of listener groups (e.g., Franken et al., 1997; Hughes et al., 2010; Hurst & Cooper, 1983). Perceived difficulties with communication often lead to academic disadvantage (e.g., Hayhow et al., 2002; Klompas & Ross, 2004; Vanryckeghem et al., 2017; Werle & Byrd, 2021), vocational role entrapment (e.g., Abou-Dahech & Gabel, 2020; Irani et al., 2009; Gabel et al., 2004; Logan & O’Conner, 2012), and exacerbates self-stigma (e.g., Boyle, 2015; Tellis & St. Louis, 2015) as well as overall poorer self-reported quality-of-life (Werle et al., 2021). Although listeners’ perception of communication competence of adults who stutter have been shown to be dissociable from stuttering severity (e.g., Werle & Byrd, 2022), limited research investigates the speaker’s perception of their own communication competence.

Self-perceived communication competence has been investigated in adolescents who stutter (Blood et al., 2001; Blood & Blood, 2004) and adults who stutter (Werle et al., 2021, Byrd et al., 2022) using the Self-Perceived Communication Competence scale (SPCC; McCroskey & McCroskey, 1998). The SPCC is a 12-item, 100-point measure of the respondents’ subjective assessment of their overall communication competence as well as their communication competence in seven specific contexts, including four different speaking scenarios (public presentation, large meeting, small group interaction, dyadic interaction) and three different interactants (strangers, acquittances, friends). As depicted in Table 1, adolescents and adults who stutter rate themselves significantly lower on the Total SPCC than non-stuttering peers, with notable variation in between-group differences on specific, context-specific subscales.

**Table 1.** Mean Scores and Standard Deviations for Respondents Who Stutter on the Self-Perceived Communication Competence Scale

|  | Blood et al. (2001) |  | Blood & Blood (2004) |  | Erickson & Block (2013)<sup>†</sup> |  | Werle et al. (2021) |  | Byrd et al. (2022)<sup>†</sup> |  |  |  |
| --- | --- | --- | --- | --- | --- | --- | --- | --- | --- | --- | --- | --- |
|  |  |  |  |  |  |  |  |  | Pre |  | Post |  |
|  | *M* | *SD* | *M* | *SD* | *M* | *SD* | *M* | *SD* | *M* | *SD* | *M* | *SD* |
| Total | 63.6* | (15.2) | 65.7* | (14.3) | 52.0 | (19.5) | 79.5* | (11.4) | 81.5 | (7.5) | 86.9 | (5.3) |
| Public | 68.2 | (9.9) | 55.4* | (8.1) | 44.5 | (27.2) | 76.4* | (14.5) | 81.1 | (8.9) | 86.3 | (5.3) |
| Meeting | 65.3 | (6.8) | 68.1 | (8.5) | 38.0 | (22.9) | 75.6 | (14.5) | 78.5 | (11.3) | 84.8 | (5.4) |
| Group | 50.7* | (9.3) | 55.1* | (9.1) | 59.2 | (22.7) | 81.5 | (10.2) | 84.0 | (6.6) | 88.9 | (5.9) |
| Dyad | 50.3* | (7.9) | 54.7* | (6.6) | 66.0 | (17.8) | 83.5 | (11.6) | 82.2 | (7.6) | 88.9 | (7.2) |
| Stranger | 49.5* | (7.0) | 55.5* | (5.9) | 38.4 | (22.4) | 72.9 | (16.5) | 71.7 | (15.3) | 81.8 | (5.8) |
| Acquaintance | 78.4 | (5.3) | 82.1 | (6.1) | 47.8 | (22.8) | 79.6* | (12.3) | 81.2 | (7.6) | 86.8 | (8.9) |
| Friend | 83.0 | (8.4) | 83.5 | (9.3) | 69.4 | (23.9) | 87.0* | (10.6) | 91.5 | (5.8) | 93.1 | (4.3) |
| *N* | 39 |  | 53 |  | 36 |  | 24 |  | 9 |  |  |  |
| Age range (y) | 13-18 |  | 13-18 |  | 11-18 |  | 18-49 |  | 18-67 |  |  |  |

\* $p < .05$ , between-group comparison with non-stuttering peers<sup>†</sup>between-group analyses not conducted because no control group was included

Additionally, as depicted in Table 2, a considerable proportion of adolescents classify themselves as “low” communication competence on the overall SPCC scale (<59 Total SPCC score; Blood et al., 2001, 41%, *n* = 39; Erickson & Block, 2013; 75%, *n* = 36) or select subscales (Blood & Blood, 2004; 45%-53%). Werle et al. (2021) found that adults who stutter also rated their communication competence significantly lower compared to age-, gender-, and education-matched fluent adults, although fewer adults than adolescents rated their communication skills in the “low” range (8%, *n* = 24) and a larger proportion rated their communication competence as “high” (>87 Total score; 21%).

**Table 2.** Percentage of Respondents who Stutter Classified as “Low” or “High” Communication Competence on the Self-Perceived Communication Competence Scale

|  | Blood et al.<br>(2001) |  | Blood & Blood<br>(2004)<sup>†</sup> |  | Erickson &<br>Block (2013) |  | Werle et al.<br>(2021) |  | Byrd et al. (2022) |  |  |  |
| --- | --- | --- | --- | --- | --- | --- | --- | --- | --- | --- | --- | --- |
|  | Low | High | Low | High | Low | High | Low | High | Pre<br>Low | High | Post<br>Low | High |
| Total | 41% | 8% | - | - | 75% | 3% | 8% | 21% | 0% | 11% | 0% | 56% |
| Public | 13% | 10% | 53% | - | 64% | 11% | 13% | 21% | 0% | 33% | 0% | 33% |
| Meeting | 15% | 8% | - | - | 75% | 3% | 17% | 30% | 0% | 22% | 0% | 44% |
| Group | 31% | 5% | 45% | - | 56% | 11% | 4% | 29% | 0% | 11% | 0% | 56% |
| Dyad | 44% | 10% | 53% | - | 61% | 3% | 4% | 38% | 0% | 11% | 0% | 44% |
| Stranger | 46% | 10% | 51% | - | 34% | 8% | 17% | 17% | 0% | 33% | 0% | 78% |
| Acquaintance | 13% | 5% | - | - | 78% | 3% | 8% | 25% | 0% | 11% | 0% | 44% |
| Friend | 10% | 10% | - | - | 47% | 11% | 0% | 28% | 0% | 22% | 0% | 11% |
| *N* | 39 |  | 53 |  | 36 |  | 24 |  | 9 |  |  |  |
| Age range (y) | 13-18 |  | 13-18 |  | 11-18 |  | 18-49 |  | 18-67 |  |  |  |

<sup>†</sup>low/high classification percentage reported only for SPCC subscales with significant between-group differences

Byrd et al. (2021) reported similarly high Total SPCC for adults who stutter pre-treatment (overall score *M* = 81.5; 0% low, 11% high) and post-treatment (Total score *M* = 86.9; 0% low, 56% high), albeit within a small, preliminary clinical trial (*n* = 9). In sum, although individuals who stutter occupy the full range of self-perceived communication competence, these ratings often remain significantly lower than non-stuttering peers. Two factors, discussed below, potentially impact relatively low self-evaluation of communication competence: treatment experience and stuttering frequency.

### Communication competence and adult stuttering treatment

Although treatments available to adults who stutter have historically sought to improve communication competence by attempting to eliminate or minimize stuttered speech (e.g., O’Brian et al., 2003; Van Riper, 1973), the benefit of focusing on fluency to improve communication competence in adults who stutter remains unclear. Neiman and Rubin (1991) found that 13 adults who stutter who completed three months of fluency-focused therapy reported increased self-perceived communication competence, as measured by the Communication Competence Self Report (Rubin, 1985), and reduced communication apprehension, as measured by the Personal Report of Communication Apprehension across speaking four contexts (dyad, group, meeting, public presentation; Daly & McCroskey, 1984). Post-treatment gains were attributed to small, yet significant, reduction in mean stuttering frequency (*M* = 21.8 pre-treatment, *SD* = 5.37; *M* = 17.5 post-treatment, *SD* = 4.75). Franken et al. (1997) investigated perceived communication *suitability* of speech samples provided by 10 adults who stutter before and after completing fluency-shaping treatment. Communication suitability was defined as sufficient situation-specific communication skills across a variety of hypothetical speaking contexts (e.g., public vs private, dyad vs group, familiar vs unfamiliar listeners) and measured from the perspective of three listener groups: naïve listeners, speech-language pathologists, and adults who stutter. Despite increased fluency among the adults who stutter, findings indicated that naïve listeners did not consider participants’ post-treatment communication abilities to be suitable across most communicative contexts, particularly for settings described as “very formal” (i.e., giving a speech or lecture, instructing a class). That is, although treatment effects were present, post-treatment speech was considered “unsuitable” or “marginally” suitable across contexts, even when stuttering-like disfluencies were comparable to control speakers (i.e., 3% stuttering-like disfluencies).

Case histories of adults who stutter suggest that previous experiences with therapy did not influence SPCC scores. For example, all 39 adolescents in Blood et al. (2001) had reported stuttering treatment within the last three years, despite significantly lower scores on the SPCC and pervasive ratings of “low” self-rated communication competence (41%). Erickson and Block (2013) reported that 84% of the 36 adolescents who stutter had undergone treatment for stuttering, with the majority of participants having completed fluency-focused treatment, despite significantly lower scores and a preponderance of self-rated “low” SPCC scores (75% of 36 adolescents who stutter). These combined data suggest that the impact of fluency-focused treatment, and perhaps fluency itself, on self- and listener-perceived communication competence for people who stutter is uncertain.

### Communication competence and stuttering frequency

A critical point to make is that perceived communication competence of adults who stutter does not predictably change based on stuttering frequency or severity (e.g., Franken et al., 1997; Gabel et al., 2008; Werle & Byrd, 2022). Werle and Byrd (2022) investigated professors’ evaluation of videos depicting an adult giving an oral presentation with 15% stuttered speech and demonstrating either high or low communication competence. Findings indicate that although stuttering frequency, content, speaker, and self-disclosure remained identical across videos, professors rated the speaker demonstrating high communication competence significantly more positively than the low communication competence sample, suggesting that stuttering and communication competence are dissociable constructs from the perspective of listeners. Werle et al. (2023) examined perceived fluency of the same video stimuli used in Werle and Byrd (2022) by untrained listeners. Results indicate that although listeners noted the stuttered speech of each video sample, produced with identical frequency and severity, listeners’ rated the stuttering of video samples with high communication competence as less distracting, and the speaker to be subjectively more fluent. Combined, findings suggest that listeners do not rely exclusively on the existence, or amount, of stuttering to determine communicative competence.

Similar to listener ratings, self-rated communication competence of people who stutter does not necessarily depend on overt stuttering behaviors. Blood et al. (2001) reported a significant, moderate correlation between stuttering severity and SPCC for adolescents who stutter. In contrast, Erickson and Block (2013) found no significant correlation between percentage of stuttered syllables and SPCC outcomes in adolescents who stutter. SPCC outcomes for adults who stutter in Werle et al. (2021) were not predicted by percentage of stuttered words, but were instead associated instead with greater adverse impact of stuttering (as measured by the Overall Assessment of Speaker’s Experience with Stuttering [OASES]; Yaruss & Quesal, 2006). Combined, these data suggest that stuttered speech is not a consistent or reliable predictor of listener- or self-perceived communication competence.

If fluency is only one of many elements considered when assessing communication competence (e.g., Spitzberg & Cupach, 2011; Spitzberg, 2007, 2013), and fluency and communication do not share a reliably significant relationship in adult who stutter (Erickson & Block, 2013; Werle et al., 2021; cf. Blood et al., 2001), one can argue that its central role in fluency-centered treatment is overemphasized. A series of studies investigating the Blank Center CARE Model^™^ approach, which explicitly excludes fluency as a clinical goal, have reported significant changes in communication competence with no shared relationship in fluency, at least from the perspective of the listener (e.g., Byrd et al., 2021; Byrd et al., 2022). Following a one-week intensive treatment program, Byrd et al. (2021) reported that clinician-rated gains in communication competence observed during oral presentations by children who stutter following treatment were not predicted by pre-treatment stuttering frequency. Byrd et al. (2022) found that pre-treatment stuttering frequency did not predict gains in clinician-rated communication competence during mock interviews by adults who stutter following 11-week CCT programming further illustrating the CCT aspect of the CARE Model. Byrd et al. (2023) recently extended these listener-based findings, reporting that untrained listeners rated post-CCT videos of an adult who stutters during a mock interview as a stronger communicator than the same adult in pre- CCT videos, despite producing equivalent stuttering severity in both videos. Combined, findings indicate that gains in communication competence for adults who stutter following CCT is reliably rated as stronger by listeners, irrespective of post-treatment stuttering severity.

Byrd et al. (2022) reported preliminary data regarding treatment effects on self-reported communication competence in adults who stutter. As noted, gains in communication competence, as rated by trained clinicians, were observed for adults who stutter during post-treatment dyadic exchanges. Findings that indicate post-treatment gains in self-perceived communication competence of people who stutter in the absence of fluency-focused clinical goals challenge previous clinical reports that attribute gains in communication competence to goals designed to conceal or minimize stuttered speech (Franken et al., 1997; Neiman & Rubin, 1991). However, although descriptive gains in SPCC were reported post-treatment by Byrd et al. (2022; pre-treatment: *M* = 81.5, *SD* = 7.5; post-treatment: *M* = 86.9, *SD* = 5.3), no significant treatment effects were obtained (*p* = .31). As noted by Byrd and colleagues, SPCC scores pre- and post-treatment were notably high (see Tables 1 and 2), and the overall sample size was relatively low (*n* = 9), and no analyses were conducted to assess the relationship between stuttering frequency and self-perceived communication competence (as measured by the SPCC). These limitations warrant further investigation of self-perceived communication competence, with a larger sample and greater range of self-perceived communication competence, as proposed in the present study.

### Purpose of the Study

Adults who stutter often report lower self-perceived communication competence than non-stuttering peers (Werle et al., 2021). Previous clinical trials examining the impact of fluency-focused treatment on self-perceived communication competence of adults who stutter have been inconclusive (e.g., Franken et al., 1997; Neiman & Rubin, 1991). Studies have yet to determine a consistent relationship between self-perceived communication competence (as measured by the SPCC) and stuttering frequency (Erickson & Block, 2013; Werle et al., 2021, cf. Blood et al., 2001). Recent clinical trials suggest that the inclusion of fluency-focused clinical goals has little to do with perceived communication competence in adolescents (Byrd et al., 2021) or adults (Byrd et al., 2023). Similar trends were identified in adults who stutter following similar communication-centered treatment (CCT; Byrd et al., 2022), but post-treatment gains based on a small sample size were non-significant. Therefore, the present study seeks to investigate the impact of CCT on adults who stutter in a larger sample of adults who stutter. Specifically, the present study will investigate the following two research questions:

**RQ1**: Does self-perceived communication competence of adults who stutter differ following CCT?

**RQ2**: Does pre-treatment stuttering frequency predict post-treatment changes in communication competence?

## Methods

The following study was approved by the authors’ university institutional review board (IRB: 2015-05-0044) and is part of an ongoing series of registered clinical trials (clinicaltrials.gov, NCT 05908123; Byrd, 2023a) designed to examine clinical outcomes of the Blank Center CARE Model^™.^ Informed consent was obtained for each adult who stutters prior to participation in data collection and treatment.

### Participants

Thirty-three adults who stutter participated in the 11-week Communication Competence Treatment (CCT) program. All participants were 18 years of age or older (*M* = 30.01, *SD* = 13.79, range: 18 to 79), self-reported proficiency in English, and either self-identified as a person who stutters and/or produced at least one stuttering-like disfluency during initial diagnostic sessions. Seventeen of the 33 participants (52%) were first-time participants at the Arthur M. Blank Center for Stuttering Education and Research. During initial diagnostic session, conducted approximately three months prior to the first treatment session, participants provided general demographic information, completed a battery of assessments designed to assess communication, fluency, cognition, language, and personal history with stuttering and stuttering treatment. Similar to Werle et al. (2021), diagnosis was confirmed via (a) a licensed speech-language pathologist analyzed three 300-word speech samples (conversational, narrative, and reading) provided by participants during diagnostic intake, (b) participant self-identified as a person who stutters, or (c) stuttering-like disfluencies and/or high impact of stuttering on life was observed during non-standardized interviews. Detailed demographic information is provided in Table 3. Information related to self-identified gender, race, ethnicity, as well as educational degree and language status were requested but not required for inclusion.

**Table 3.**
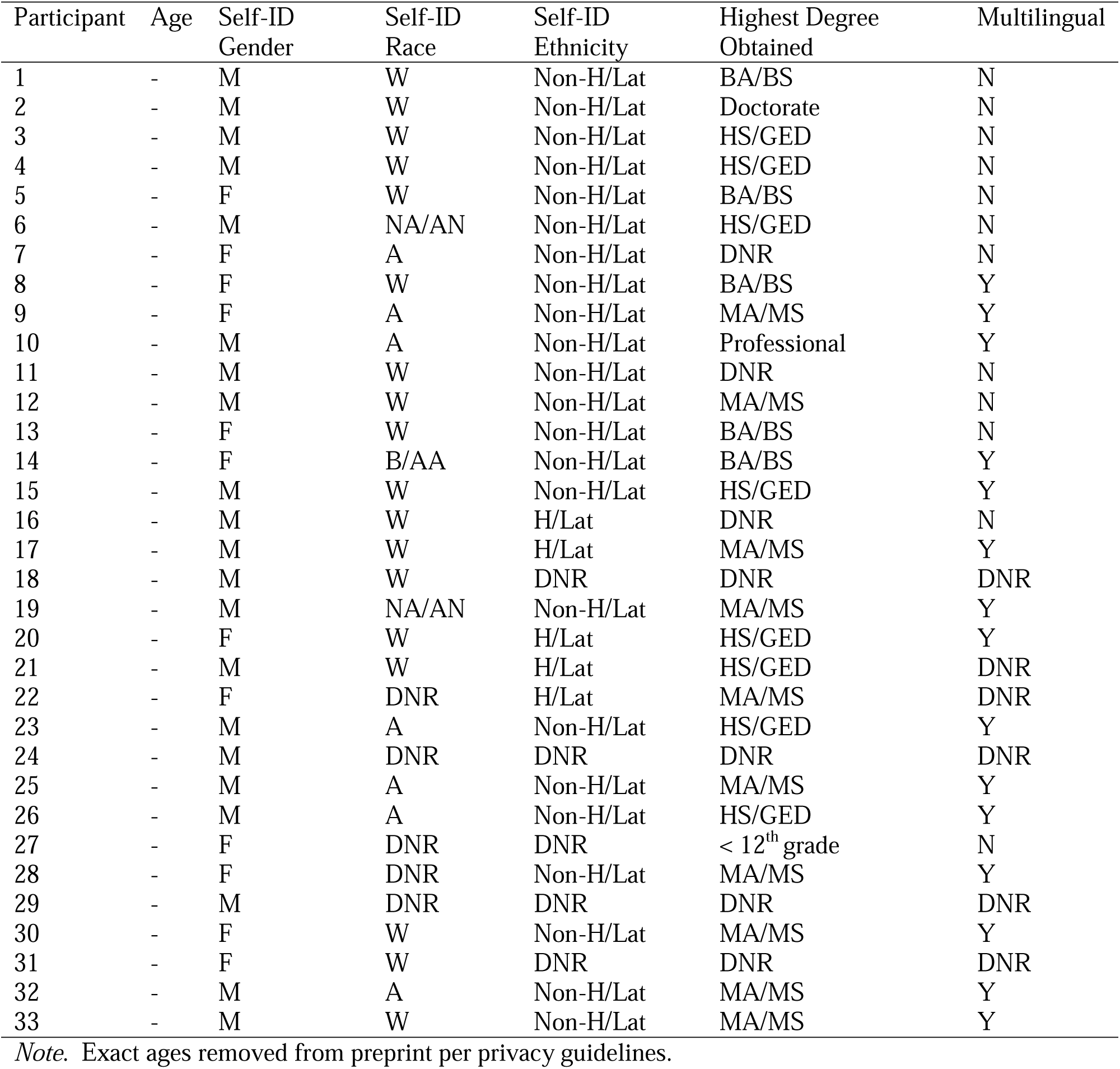
Demographic information of participants

| Participant | Age | Self-ID<br>Gender | Self-ID<br>Race | Self-ID<br>Ethnicity | Highest Degree<br>Obtained | Multilingual |
| --- | --- | --- | --- | --- | --- | --- |
| 1 | - | M | W | Non-H/Lat | BA/BS | N |
| 2 | - | M | W | Non-H/Lat | Doctorate | N |
| 3 | - | M | W | Non-H/Lat | HS/GED | N |
| 4 | - | M | W | Non-H/Lat | HS/GED | N |
| 5 | - | F | W | Non-H/Lat | BA/BS | N |
| 6 | - | M | NA/AN | Non-H/Lat | HS/GED | N |
| 7 | - | F | A | Non-H/Lat | DNR | N |
| 8 | - | F | W | Non-H/Lat | BA/BS | Y |
| 9 | - | F | A | Non-H/Lat | MA/MS | Y |
| 10 | - | M | A | Non-H/Lat | Professional | Y |
| 11 | - | M | W | Non-H/Lat | DNR | N |
| 12 | - | M | W | Non-H/Lat | MA/MS | N |
| 13 | - | F | W | Non-H/Lat | BA/BS | N |
| 14 | - | F | B/AA | Non-H/Lat | BA/BS | Y |
| 15 | - | M | W | Non-H/Lat | HS/GED | Y |
| 16 | - | M | W | H/Lat | DNR | N |
| 17 | - | M | W | H/Lat | MA/MS | Y |
| 18 | - | M | W | DNR | DNR | DNR |
| 19 | - | M | NA/AN | Non-H/Lat | MA/MS | Y |
| 20 | - | F | W | H/Lat | HS/GED | Y |
| 21 | - | M | W | H/Lat | HS/GED | DNR |
| 22 | - | F | DNR | H/Lat | MA/MS | DNR |
| 23 | - | M | A | Non-H/Lat | HS/GED | Y |
| 24 | - | M | DNR | DNR | DNR | DNR |
| 25 | - | M | A | Non-H/Lat | MA/MS | Y |
| 26 | - | M | A | Non-H/Lat | HS/GED | Y |
| 27 | - | F | DNR | DNR | < 12<sup>th</sup> grade | N |
| 28 | - | F | DNR | Non-H/Lat | MA/MS | Y |
| 29 | - | M | DNR | DNR | DNR | DNR |
| 30 | - | F | W | Non-H/Lat | MA/MS | Y |
| 31 | - | F | W | DNR | DNR | DNR |
| 32 | - | M | A | Non-H/Lat | MA/MS | Y |
| 33 | - | M | W | Non-H/Lat | MA/MS | Y |

*Note.* Exact ages removed from preprint per privacy guidelines.

### Communication Competence Treatment (CCT)

CCT is an 11-week treatment program comprised of two weekly sessions (one group session, one individual session). The 22-session CCT approach is informed by the speaker’s experience with stuttering, and reflective of contemporary definitions of this complex condition (for manual, see Byrd, 2023b; also see Byrd, 2021; Byrd et al., 2016; Constantino, 2018; Watermeyer & Kathard, 2016; Usler, 2022). Unlike fluency-centered approaches that are rooted in ableism (i.e., fixing a disabling condition), and impose societal pressures to conform to speaking fluently, CCT facilitates authenticity, and spontaneity in the communication exchanges of those who stutter (Byrd et al., 2021; Byrd et al., 2022; Byrd et al., 2023; Young et al., 2022). Participants learn competencies that are core to effective communication for all speakers (e.g., language organization, nonverbals, pragmatics, vocal emphasis; Morreale et al., 2007; Spitzberg, 2007). They also learn competencies that support positive experiences with stuttering (e.g., mindfulness, acceptance and commitment, self-compassion, self-disclosure) with the goal of using these skills across a variety of functional yet challenging environments, irrespective of fluency. Thus, CCT increases communication competency, promotes agency, and rejects the negative assumptions associated with stuttering as a disability (Byrd et al., 2022a; Croft & Byrd, 2020; Byrd & Croft, in press; Kwon, 2021; Werle & Byrd, 2022).

Byrd (2023b) details the manualized 22-session CCT program developed as part of the Blank Center CARE Model^™^ (Communication, Advocacy, Resilience, and Education). In brief, treatment consists of two 60-minute sessions per week, including explicit instruction in (1) effective communication skills (e.g., language organization, nonverbals, pragmatics, vocal emphasis) in a number of challenging formal and spontaneous communicative exchanges (e.g., icebreakers, small group presentations, large group presentations, one-on-one mingling, mock interviews, impromptu dyadic interactions and presentations; Byrd et al., 2021; 2022; 2023), (2) advocacy skills, such as self-disclosure (Byrd et al., 2017; Croft & Byrd, 2021; Werle & Byrd, 2022) and navigating negative peer responses, (3) resilience training, including mindfulness and self-compassion (Croft & Byrd, 2020; Byrd & Croft, in press; Harris, 2019), and (4) education about stuttering, including the basic facts and misperceptions/stereotypes (Byrd et al., 2017; Werle & Byrd, 2022). Participants culminate their CCT program by presenting a reflective summary via a presentation for a larger audience of > 200 familiar and unfamiliar people.

### Self-Perceived Communication Competence scale (SPCC; McCroskey & McCroskey, 1988)

The Self-Perceived Communication Competence (SPCC; McCroskey & McCroskey, 1988) is a brief 12-item scale designed to assess an individual’s perception of their communication competence across four specific communicative contexts (dyad, small group, large meeting, presentation) and three interlocutors (stranger, friend, acquaintance). The SPCC generates one composite scale ratings (Total SPCC) and seven subscale ratings - four subscales categorized by speaking context (i.e., public presentation, large meeting, small group interaction, dyadic interaction) and three subscales categorized by audience (i.e., stranger, acquaintance, friends). Higher scores on a 100-point scale (0 = completely incompetent, 100 = completely competent) reflect stronger communication competence. Total scores above 87 are indicative of high self-perceived communication competence and scores below 59 are indicative of low self-perceived communication competence. The original study, based on a sample of college students in the United States, reported an excellent reliability (Cα ≥ .92) for the total score and test-retest reliability of .77 to .89. Unlike most self-rated measures of communication, and consistent with the nature of stuttering and the Blank Center CARE Model^™^, the SPCC incorporates and isolates communication competence in different speaking scenarios rather than focusing on interpersonal conversation alone.

### Stuttering frequency

Pre-treatment stuttering frequency was calculated based on impromptu presentations delivered by participants during the first week of CCT. Stuttering-like disfluencies were classified as sound and syllable repetition, audible prolongations, and/or inaudible prolongations produced with atypical stress and/or rhythm. Videotaped presentations were coded offline by a licensed, certified speech-language pathologist (Coder 1) and an undergraduate student clinician (Coder 2), both of whom were trained in disfluency count protocol and familiar with the Blank Center CARE Model^™^, yet unfamiliar with the participants depicted in each video. Each coder rated approximately half of the cohort (Coder, n = 15; Coder 2, n = 18). To determine inter-rater reliability, Coder 1 analyzed approximately one-third of samples completed by Coder 2 (n = 6 of 18, 33%), and vice versa (n = 5 of 15, 33%). To determine intra-rater reliability, Coders 1 and 2 re-analyzed one-third of their own cohort (Coder 1, n = 6 of 18, 33%; Coder 2, n = 5 of 15, 33%) at least four weeks after initial completion without knowledge of previous scores. Intra-rater reliability was sufficiently high (*r* = .98, ICC = .99; *p* < .001) as was inter-rater reliability (*r* = .83; inter-item correlation coefficient [ICC] = .88; *p* < .001).

### Data analyses

To examine RQ1, paired t tests were conducted to compare self-perceived communication competence pre- and post-treatment for adults who stutter. Bonferroni-Holm corrected *p*-values (Holm, 1979) were applied to accommodate multiple comparisons across Total SPCC scale and seven SPCC subscales (public, meeting, group, dyad, stranger, acquaintance, friend). Significant pre-/post-treatment changes identified during parametric t- tests were confirmed by nonparametric Wilcoxon’s signed-rank test. Effect sizes were calculated using Cohen’s *d* (Cohen, 1998; .2 = small, .5 medium, .8 large). To examine RQ2, eight separate linear regressions were conducted with pre-treatment stuttering frequency as the sole predictor, and differences between self-perceived communication competence pre- to post-treatment as measured by the Total SPCC and each of the seven SPCC subscales.

## Results

**RQ1**: Does self-perceived communication competence of adults who stutter differ following CCT?

As depicted in Figure 1 and Table 4, paired t-test analyses indicate significant gains in self-perceived communication competence post-treatment compared to pre-treatment based on the composite SPCC scale *t*(32) = 4.12, *p* < .0001, *d* = .72 [medium-large effect size].

**Figure 1.**
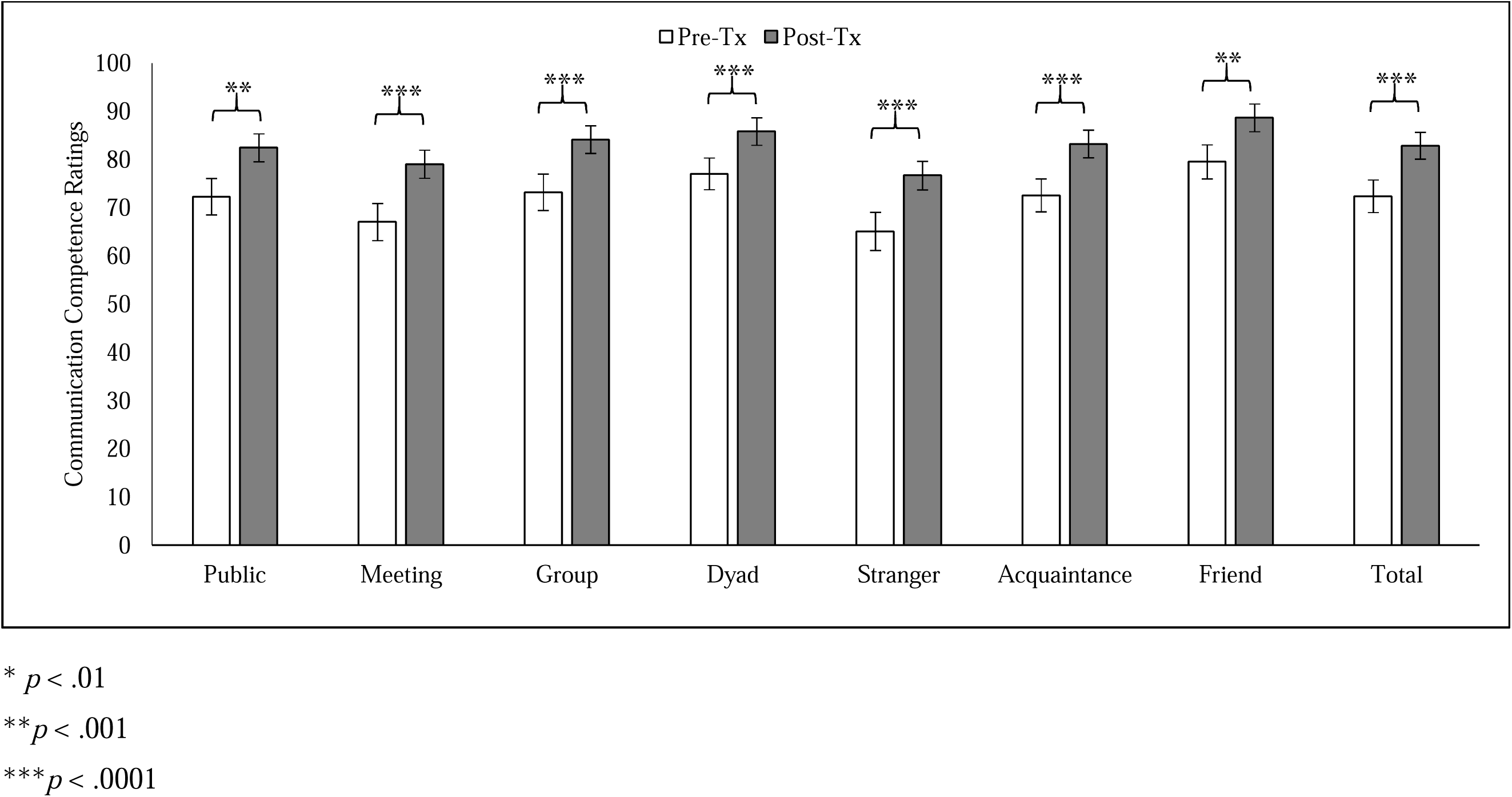
Self-perceived communication competence reported by adults who stutter before treatment (Pre-Tx) and after treatment (Post-Tx) across speaking situations included in the SPCC (McCroskey & McCroskey, 1988).

**Table 4.**
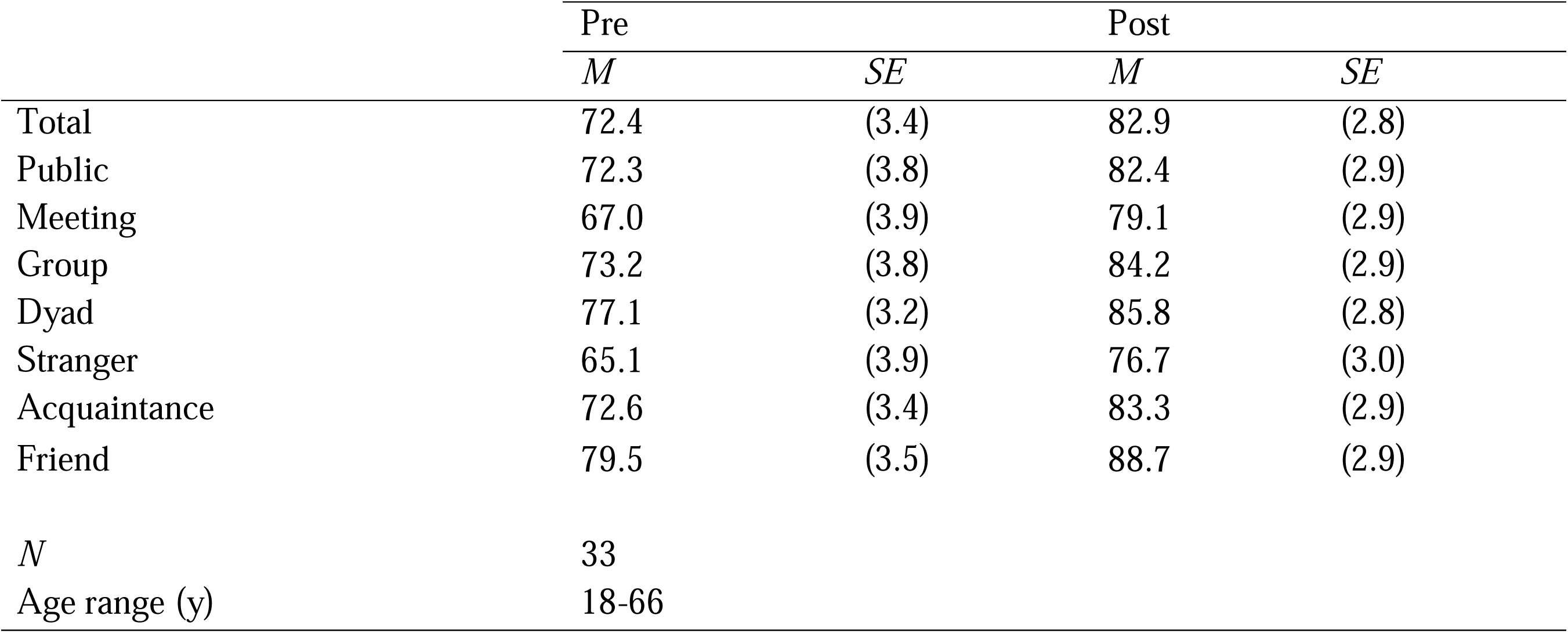
SPCC Scores (Mean, Standard Error of the Mean) for Adults Who Stutter Pre- and Post- Treatment

Significant gains were observed for all four speaking situations, including public presentation *t*(32) = 3.49, *p* < .001, *d* = .61 [medium-large effect size], large meeting *t*(32) = 3.71, *p* < .0001, *d* = .65 [medium-large effect size], small group *t*(32) = 3.54, *p* < .001, *d* = .62 [medium-large effect size], and dyadic interaction *t*(32) = 3.94, *p* < .0001, *d* = .69 [medium-large effect size].

Significant gains were also reported for all three audience types, including stranger *t*(32) = 3.86, *p* < .0001, *d* = .67 [medium-large effect size], acquaintance *t*(32) = 4.07, *p* < .0001, *d* = .71 [medium-large effect size], and friend *t*(32) = 3.71, *p* < .001, *d* = .61 [medium-large effect size].

Significant gains were maintained for all subscales after Bonferroni-Holm p-value adjustment for multiple comparisons (Bonferroni-Holm adjusted *p*-value range: .002 to .004). Significant gains for all subscales were further verified via nonparametric analyses (*p*-value range: .005 to .002). Table 5 indicates the proportion of adults who stutter who reported high/low communication competence, per the SPCC, pre- and post-treatment.

**Table 5.** Percentage of Adults who Stutter Classified as “Low” or “High” Communication Competence on the SPCC Pre- and Post-Treatment

|  | Pre |  | Post |  |
| --- | --- | --- | --- | --- |
|  | Low | High | Low | High |
| Total | 21% | 21% | 3% | 42% |
| Public | 12% | 30% | 0% | 48% |
| Meeting | 24% | 24% | 3% | 42% |
| Group | 24% | 18% | 3% | 52% |
| Dyad | 24% | 21% | 3% | 36% |
| Stranger | 9% | 24% | 0% | 61% |
| Acquaintance | 27% | 15% | 3% | 30% |
| Friend | 33% | 21% | 6% | 21% |
| *N* | 33 |  |  |  |
| Age range (y) | 18-66 |  |  |  |

**RQ2**: Does pre-treatment stuttering frequency predict post-treatment changes in communication competence?

As summarized in Table 6, pre-treatment stuttering frequency did not predict post-treatment gains in Total SPCC (*p* = .62) or any of the seven SPCC subscales (*p*-value range: .26 to .96)

**Table 6.** Eight simple linear regression models with self-perceived communication competence as the dependent variable (SPCC Total and seven SPCC subscales) and stuttering frequency as the sole

| | B | $\beta$ | p |
| --- | --- | --- | --- |
| Public | .15 | .06 | .74 |
| Meeting | .56 | .20 | .26 |
| Group | .02 | .01 | .96 |
| Dyad | .03 | .34 | .93 |
| Stranger | .42 | .17 | .36 |
| Acquaintance | .29 | .13 | .48 |
| Friend | -.14 | -.06 | .73 |
| Total | .19 | .09 | .62 |

## Discussion

The present study investigated the outcomes of self-perceived communication competence amongst adults who stutter following communication-centered treatment (CCT).

Given the inconsistent relationship in previous literature between self-perceived communication competence and frequency or severity of stuttering, this study also sought to investigate whether outcomes were predicted by stuttering frequency. Findings indicate significant, positive post-treatment gains in self-perceived communication competence, as measured by the SPCC, for adults who stutter irrespective of pre-treatment stuttering frequency. Results of the present study add to the growing body of literature investigating the positive cognitive, affective, and communication outcomes of the Blank Center CARE Model^™^.

**RQ1**: Does self-perceived communication competence of adults who stutter differ following CCT?

Following participation in the 11-week CCT programming, participants demonstrated significant increases in their perceived competence in communicating across each of the seven sub-domains (4 speaking contexts, 3 audience types) measured by the SPCC, also reflected in the composite scores. These findings extend the preliminary data reported by Byrd et al. (2022) which found descriptive, but not statistically significant, improvements in SPCC scores for a smaller sample of adults who stutter (n = 9) following participation in CCT. As depicted in Table 5, participants in the current study also exhibited lower levels of self-perceived communication competence prior to treatment (i.e., 21% of 33 participants reported ‘low’ Total Scores) compared to Byrd et al. (2022; 0% of 9 participants reported ‘low’ Total Scores) on SPCC. The relatively normal distribution of scores pre-treatment (21% low, 58% typical, 21% high) from a larger cohort of adult participants provide confidence that positive trends observed in Byrd et al. (2022) were not due to smaller clinical cohort with negatively skewed (i.e., higher) SPCC pre-treatment scores. Findings extend previous studies by Byrd et al (2022) and Byrd et al. (2023) by documenting significant gains in communication abilities, this time from the perspective of the client, achieved by adults who stutter following CCT across a variety of communication contexts.

As summarized in Table 4, the adult in the present study reported Total Scores at pre-treatment (*M* = 72.4) that were higher than previous studies investigating the self-perception of communication abilities in adolescents who stutter (Blood et al., 2001 [*M* = 63.6]; Blood & Blood, 2004 [*M* = 65.7]) and considerably higher than adolescents who stutter sampled by Erickson and Block (2013; *M* = 52.0). Patterns that suggest greater confidence with communicative abilities with age are consistent with previous publications that have reported self-perceived communication abilities may evolve over time, and with exposure to a variety of speaking situations, for both non-stuttering adults (Kiessling & Fabry, 2021) and adults who stutter (Werle et al., 2021). Total Scores in the present study were slightly lower, however, than adults who stutter investigated in Werle et al. (2021, *M* = 79.5, *n* = 24), who in turn reported significantly lower SPCC than non-stuttering peers (*M* = 86.0, *n* = 26). Thus, although the present sample of adults who stutter were not report disproportionately low or high communication competence, pre-treatment scores may reflect more negative self-appraisal than age-, gender-, and education-matched peers who do not stutter documented in Werle et al. (2021). Although post-treatment SPCC scores in the present study (M = 82.9) were comparable to non-stuttering adults in Werle et al. (2021), further clinical trials with non-stuttering control groups would be necessary to verify this claim. It is also important to note that the differences observed across cohorts in the current and previous publications indicates that individuals who stutter exist along a continuum with respect to their self-perceptions of their skills as communicators.

Similar to Byrd et al. (2022), participants of the present cohort included both individuals who were engaging in treatment for the first time (n = 16, 49.5%) as well as individuals who were returning, or had completed at least one iteration of the 11-week protocol previously (n = 17, 51.5%). Unlike Byrd et al. (2022, n = 11, four new participants, seven returning participants), the relatively even distribution of new clients and returning clients suggests that significant post-treatment gains were not limited to those who were experiencing CCT for the first time. Further post-hoc analyses of Total SPCC Scores for each group confirm that both new clients (*t*(16) = 3.23, p = .005) and returning clients (*t*(15) = 3.50, *p* = .030) reported significant gains post-treatment, despite smaller sample sizes. Although additional clinical trials are warranted to determine whether specific aspects of CCT are more beneficial to new participants versus returning participants, combined findings demonstrate the benefit of the clinical program to self-perceived communication competence for adults who stutter from the perspective of clinicians (Byrd et al., 2022) and clients (current study), irrespective of prior engagement with CCT.

**RQ2**: Does pre-treatment stuttering frequency predict post-treatment changes in communication competence?

Stuttering frequency, as measured by percentage of stuttered syllables, did not predict any of the changes to SPCC scores across any of the seven subscales nor the Total Score following CCT treatment. These results align with previous research studies which have documented no linear relationship between stuttering frequency and SPCC scores (e.g., Erickson & Block, 2013; Werle et al., 2021). Although Blood et al. (2001) did find a moderate relationship between the SPCC and stuttering severity, as measured by the SSI-4, it is possible that this relationship was driven by the additional parameters included on the SSI-4 (e.g., secondary behaviors, temporal duration of stuttering moments). As noted by Werle et al. (2021), SPCC scores were driven by greater adverse impact of stuttering (as measured by the OASES) than stuttering severity.

Although Blood et al. (2001) did not include quality-of-life measures such as the OASES, given that their cohort presented with relatively high levels of communication apprehension and low self-perceived communication competence, it is possible that the relationship between the SSI-4 score and the SPCC was more driven by participants’ negative communication attitude rather than stuttering frequency. It is also possible, given the growing number of studies that have found no consistent relationship between stuttering severity and communication competence, that the presumed relationship between these factors has been overestimated.

Previous studies specifically examining CCT have also found that improved communication abilities have not been predicted by stuttering frequency, both for adults who stutter (e.g., Byrd et al., 2022; Byrd et al., 2023, present study) and adolescents (Byrd et al., 2021). Findings from the present study also provide counterevidence to Neiman and Rubin (1991), who reported significantly higher communication competence for 13 adults who stutter following fluency-focused treatment that was accompanied by significant reduction in stuttering severity post-treatment. To reiterate, fluency was neither the focus nor the desired outcome of CCT, and incidental changes in fluency post-treatment were just that – incidental. Thus, similar to Byrd et al. (2021, 2022), the current study did not analyze post-treatment stuttering severity in relation to post-treatment communication competence. To measure post-treatment fluency would only reinforce the concept that communication abilities should always be qualified or measured within the lens of stuttering severity. Instead, our findings provide evidence for a treatment approach to that challenges the common assumption that communication competence in adults who stutter requires speakers to conceal, or attempt to conceal, stuttered speech. Such gains in self-perceived communication competence with no attempt to minimize stuttered speech undermine the notion targeting fluency – a clinical outcome that is difficult to maintain (e.g., Cream et al., 2003; NSA, 2009) and considered by many adults who stutter to be part of an identity rather than a “disorder” (e.g., Byrd, 2021; Constantino, 2018) – as an effective or ethical approach to stuttering treatment.

### Clinical Implications

The outcomes of the present study have significant implications for individuals who stutter, allies, and professionals who support them. Improvement in self-perception of communication abilities are particularly critical to facilitate amongst individuals who stutter, especially those who fall within ‘low’ cutoff ranges on the SPCC. Functionally, many communication behaviors are dictated by one’s self-perception of communication abilities (Phillips, 1984). Individuals who have poor self-appraisals of communication skills may be more reticent to communicate and thus engage in their everyday environments, leading to potential isolation and reduced quality of life (Blood et al., 2021; Werle et al., 2021). Individuals who stutter are vulnerable to such negative experiences, whether due to low self-appraisal of communication abilities, societal reactions to stuttering, or other environmental factors (Blood & Blood, 2016; Constantino et al., 2022). The CCT protocol implemented in the present study has preliminary evidence that, in addition to improving self-perception of communication abilities, also improves observer-appraisal of effective communication skills, as well as increased resilience and a reduction in the negative influence of stuttering on overall quality of life (Byrd et al., 2022; Byrd et al., 2023).

### Limitations and Future Research

Findings from the present study should be interpreted with the following considerations in mind. First, the study sample included 33 adults who stutter. Although currently the largest clinical trial to examine self-perceived communication competence in individuals who stutter, future clinical trials should attempt to recruit a larger sample size to further determine CCT efficacy. Additionally, the present study investigated the influence of one clinical variable, stuttering frequency, on post-treatment SPCC scores. A larger sample size would enable exploration of additional clinical and psychosocial predictors of self-perceived communication competence, such as self-reported participation in communication situations in daily life, frequency and use of self-disclosure, self-compassion, and/or self-stigma. Given the potential relationship between these variables and self-perceived communication competence (Boyle, 2013; Boyle, 2015; Byrd et al., 2022; Croft & Byrd, 2020; Werle & Byrd, 2022; Young et al., 2022), clarifying the relative contribution of each would allow for more precise and effective clinical intervention.

The present study used a pre- to post-test design to determine the influence of CCT on self-perceived competence. A control group was not employed, and participants’ activities outside of CCT over the course of treatment were not controlled, nor was a longitudinal design employed to investigate long-term stability of post-treatment outcomes. Future studies would benefit from both to isolate the influence of CCT treatment on observed outcomes, perhaps by including a waitlist control group and monitoring participants’ activities outside of the treatment program, such as concurrent enrollment in other communication-related activities.

It should also be noted that participants engaged in 22 treatment sessions with a variety of activities and topics. It is unknown which aspects of CCT are most effective as it relates to improving self-perceived communication competence. It is possible that certain activities are more predictive of improvement in self-perceived communication competence than others, and that these significant predictors differ depending on an individual’s unique clinical and psychosocial profile. Thus, future studies could explore the relative impact of key CCT components to determine which activities are most beneficial and for whom. Studies could also utilize a qualitative approach to identify participants’ perspectives on CCT and to elucidate the functional impact of CCT on participants’ daily lives, including the most effective or beneficial aspects of the program.

Although statistically significant improvements in self-perceived communication competence were reported at post-treatment, as measured by the SPCC scores, participants’ communication competence across communication contexts and partners (i.e., friend, acquaintance, stranger) was not rated by a third-party. Thus, the precise relationship between self- and observer-rated communication competence across each communication context and partner, as measured on the SPCC, is unknown and denotes an area for future study. Taken together, these limitations provide direction for future research to broaden the application of study findings to the population of adults who stutter.

## Conclusion

Findings demonstrate the benefits of CCT to self-rated communication competence, as measured by the SPCC, for adults who stutter across a variety of speaking situations and audience. Results corroborate outcomes from previous clinical trials from the perspective of clients, wherein significant communication gains were observed from the perspective of naïve observers and speech-language pathologists. Findings provide further evidence that gains in communication competence for individuals who stutter are observed in the absence of clinical goals targeting fluency.

## Data Availability

All data produced in the present study are available upon reasonable request to the authors

## Data Availability

All data produced in the present study are available upon reasonable request to the authors

## Acknowledgements

This project was supported by the foundational grant support funded to the Arthur M. Blank Center for Stuttering Education and Research and endowed support provided through the Michael and Tami Lang Stuttering Institute, the Dr. Jennifer and Emanuel Bodner Developmental Stuttering Laboratory, and the Dealey Family Foundation Stuttering Clinic awarded to the second author. The authors would like to thank Caitlin Franchini, Diana Perez, Hudson Hanna, and Zaniel Flores for reliability coding, as well as JoLynn Riojas for video preparation efforts. We would also like to thank the adult and child participants who stutter, as well as their families, who continue to participate in our ongoing clinical research.

## Notes

### Competing Interest Statement

The authors have declared no competing interest.

### Clinical Trial

NCT05908123

### Funding Statement

This study did not receive any funding

### Author Declarations

The study was approved by the institutional review board (IRB: 2015-05-0044) at the University of Texas at Austin.

